# A comparative evaluation of a dye-based and probe-based RT-qPCR assay for the screening of SARS-CoV-2 using individual and pooled-sample testing

**DOI:** 10.1101/2020.05.30.20117721

**Authors:** Claudio Verdugo, Anita Plaza, Gerardo Acosta-Jamett, Natalia Castro, Josefina Gutiérrez, Carlos Hernández, Carmen López-Joven, Carlos A. Loncoman, Claudio Navarrete, Alfredo Ramírez-Reveco, Alex Romero, Andrea Silva, Matías Vega, Cristóbal Verdugo, Jonathan Vergara-Amado

## Abstract

Effective interventions are mandatory to control the transmission and spread of SARS-CoV-2, a highly contagious virus causing devastating effects worldwide. Cost-effective approaches are pivotal tools required to increase the detection rates and escalate further in massive surveillance programs, especially in countries with limited resources that most of the efforts have focused on symptomatic cases only. Here, we compared the performance of the RT-qPCR using an intercalating dye with the probe-based assay. Then, we tested and compared these two RT-qPCR chemistries in different pooling systems: after RNA extraction (post-RNA extraction) and before RNA extraction (pre-RNA extraction) optimizing by pool size and template volume. We evaluated these approaches in 610 clinical samples. Our results show that the dye-based technique has a high analytical sensitivity similar to the probe-based detection assay used worldwide. Further, this assay may also be applicable in testing by pool systems post-RNA extraction up to 20 samples. However, the most efficient system for massive surveillance, the pre-RNA extraction pooling approach, was obtained with the probe-based assay in test up to 10 samples adding 13.5 µL of RNA template. The low cost and the potential use in pre-RNA extraction pool systems, place of this assays as a valuable resource for scalable sampling to larger populations. Implementing a pool system for population sampling results in an important savings of laboratory resources and time, which are two key factors during an epidemic outbreak. Using the pooling approaches evaluated here, we are confident that it can be used as a valid alternative assay for the detection of SARS-CoV-2 in human samples.

## INTRODUCTION

The Severe Acute Respiratory Syndrome Coronavirus-2 (SARS-CoV-2) is the etiological agent of the globally spread Coronavirus Disease-2019 (COVID-19). The disease was first reported in late December 2019 in Wuhan, China. By the time the WHO declared the COVID-19 outbreak a pandemic (March 12th), there were around 1,000 deaths over ∼20,000 confirmed cases. Up to date (May 30th), more than 350,000 individuals have died and 6 million cases have been confirmed (WHO, 2020a). Genome analysis approach has been essential to tackle the COVID-19 pandemic from the beginning as the SARS-CoV-2 nucleotide identity was rapidly uploaded to public databases, resulting in the immediate manufacture of quantitative reverse transcription PCR (RT-qPCR) tests (Ren et al., 2020). Further, genome analysis helped to highlight notable genomic features of SARS-CoV-2 (Andersen et al., 2020) that in agreement with structural and biochemical experiments determined that the glycoprotein S was optimized for binding the human receptor hACE2 and that such increase in binding affinity in SARS-CoV and SARS-CoV-2 was correlated with an increase in virus pathogenicity (Li et al., 2005; Walls et al., 2020). These similarities among the SARS-CoV-2 and SARS-CoV indicates that differences in the epidemiology of COVID-19 and SARS, respectively, probably arise from other factors, including high viral loads in the upper respiratory tract and the potential for asymptomatic carriers (Bai et al., 2020; He et al., 2020; Liu et al., 2020).

Asymptomatic and pre-symptomatic individuals are one of the most important keys to control the transmission and spread of SARS-CoV-2. According to recent reports, up to 80% of infections are mild or asymptomatic, whereas a 15% develop severe infection and 5% are critical infections (CDC, 2020; Bai et al., 2020; Li et al., 2020; Mizumoto et al., 2020; WHO, 2020b). Further, a significant portion of transmissions occurs before infected patients develop symptoms (He et al., 2020), and a drastic underestimation of COVID-19 case number by up to 50 to 100-fold has been suggested (Richterich, 2020). Specifically, viral shedding starts around2.3 days and peaks ∼1 day before the onset of the symptoms. Hence, finding the proportion of pre-symptomatic and asymptomatic transmission is mandatory to establish an active case finding and further control of the disease. A case identification followed by tracing and quarantining contacts and high-risk personnel are useful approaches when a new infection is emerging in a population (Hellewell et al., 2020; Wu and McGoogan, 2020). However, when a community transmission is already established, tracing the contact is an incomplete method for controlling transmission. Thus, the cases detected through those methods are no longer representative of the whole infection dynamic on the population, limiting the capacity to make evidence-based decisions (Foddai et al., 2020). For those reasons on the current scenario, urge to massively increase the diagnostic rates allowing to identify not only symptomatic cases but, most importantly, asymptomatic and pre-symptomatic cases (Peto, 2020; Peto et al., 2020).

The early identification of individuals undergoing viral shedding has evident epidemiological, economic, and social benefits (e.g. Pareek et al., 2016; Prins et al., 2017). However, in developing countries, implementing a massive testing approach remain still unpracticable. The official confirmatory test is a probe-based RT-qPCR (CDC, 2020). The current pandemic situation has created a shortage of reagents, limiting the use for individual and confirmatory testing only. In consequence, alternative detection assays, such as the use of intercalating dyes, and different analytical techniques, such as pooling testing, that may retain the diagnostic sensitivity and specificity of the current official and individual assay is pivotal to progress for massive use. Furthermore, given the expected low prevalence of SARS-CoV-2 in the general population, massive testing would render mostly negative results. A sampling strategy based on the pooled analyses, instead of individual samples, could be an efficient and effective approach to reduce cost and enlarge the number of tested individuals for massive screening in the general population, notably reducing the assay cost (Eis-Huebinger et al., 2020; Narayanan et al., 2020). The fluorescent DNA-intercalating dye-based technology is alternative chemistry for RT-qPCR assay due to several factors such as low cost, feasible to perform, and highly scalable compared to probes-based assays (i.e. TaqMan probes) (Tajadini et al., 2014). Therefore, the present study aimed to evaluate the sensitivity and performance of two RT-qPCR assays, a dye-based and a probe-based, for the detection of SARS-CoV-2 in human samples. Then, we compared the performance of both assays by pooling samples at different group sizes and RNA template volume. We further evaluated pooling samples at two different analytical stages, preand post-RNA extraction.

## MATERIAL AND METHODS

### Ethics Statement

Samples were obtained from diagnostic certified laboratory and collected during two-weeks in a surveillance program of risk group of health care workers and military personnel. Ethical approval of this study was obtained from the Comité Ético Científico – Servicio de Salud Valdivia (CEC-SSV, reference number Ord. Nº099).

### Clinical Samples and RNA Extraction

A total of 610 nasopharyngeal samples were obtained using swabs (FLOQSwabs #503, COPAN Diagnostic Inc., Italy) and deposited in 2 mL universal viral transport media. Viral RNA was extracted from 200 µL of the transport media containing the sample using Viral Nucleic Acid Extraction Kit II (GeneAid) according to manufacturer’s instructions, obtaining a final elution of 60 µL with nuclease-free water. The quality of the RNA and the RT-qPCR assay were evaluated by amplifying the human *RNAseP* gene as an internal control. An extraction control was performed in each extraction run. All real-time RT-PCR reactions were done with an Applied Biosystems QuantStudio3 Real-Time PCR System (Thermo Scientific).

### Dye-based RT-qPCR

A single-tube one-step RT-qPCR was standardized using a GoTaq® 1-Step RT-qPCR System (Promega). This kit contains the fluorescent BRYT Green® dye wich has similar spectral properties to SYBR® Green I, such as excitation at 493 nm and emission at 530 nm. A set of three pairs of primers were synthesized targeting different segments of the nucleocapsid (Table 1). Primer concentration (0.2 to 1.5 µM) and annealing temperature (58 to 65ºC) were optimized to achieve a better performance. A concentration of 0.3 µM of each primer and 62.5ºC was found to yield the lowest threshold cycles (C_T_ values), highest fluorescence signal, and minimal secondary products. For individual samples, RT-qPCR was performed with a final volume of 20 µL containing 10 µL of master mix, 5 µL of RNA template, 0.6 µL of each primer (10 µM), and 3.8 µL of nuclease-free water. The optimized cycling profile was 50°C for 15 min, 95°C for 10 min, followed by 40 cycles of 95°C for 15 sec, 62.5°C for 15 sec, and 72ºC for 15 sec. Melting analysis was performed by 15 sec at 95°C and then an incubation of 1 min at 60°C with a gradual increase in temperature (0.15°C /1s) to 95°C. The fluorescence was measured at the end of each cycle. A human specimen control (i.e. RNAseP gene), a synthetic viral nucleocapside positive control (2019-CoV Plasmid Control, IDT), and a negative control (ultra-pure water) were included during each RT-qPCR run.

**Table 1.**
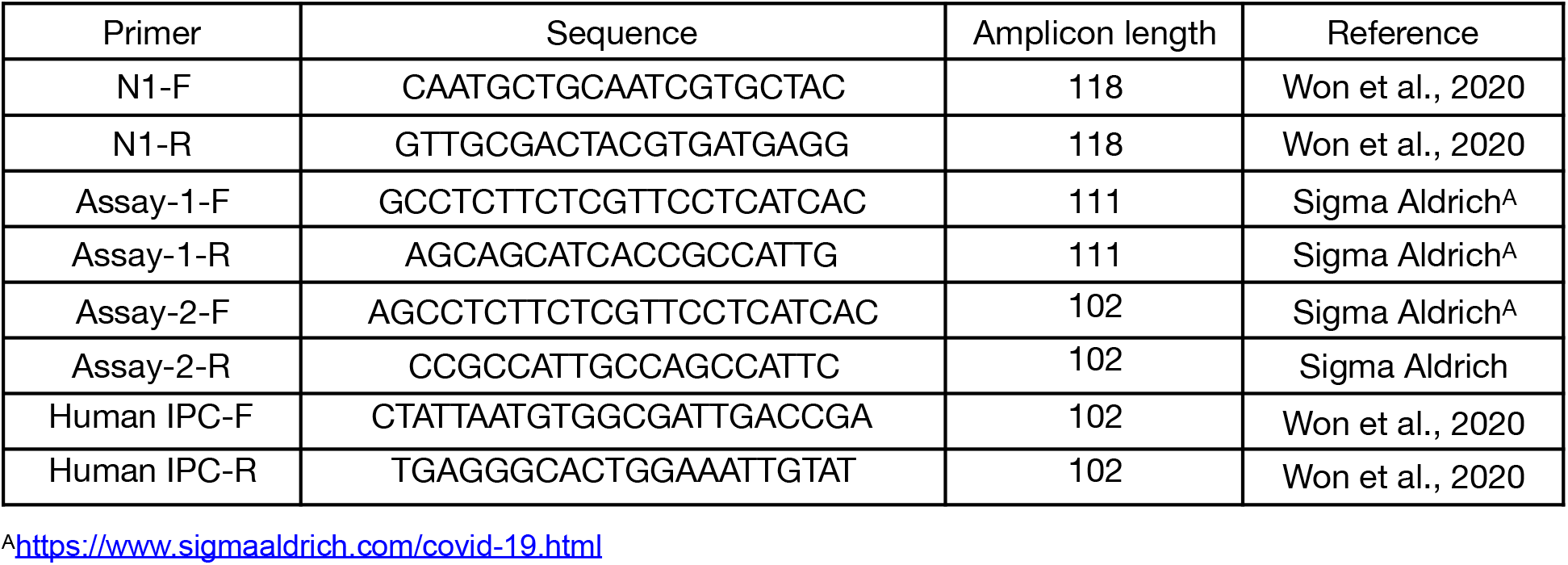
Primers for nucleocapside amplification of SARS-CoV-2 by a dye-based assay used in this study.

### Probe-based RT-qPCR

Two segments of the nucleocapsid gene (i.e. N1 and N2) were amplified using 4x TaqMan® Fast Virus 1-step Master Mix (Thermo Fisher) in a 20 µL reaction containing 5 µL of master mix, 5 µL of RNA template, 1.5 µL of primer and probe mix (2019-nCOV CDC EUA Kit, IDT), and 8.5 µL of nuclease-free water. Briefly, the cycling profile was 25ºC for 2 min, 50ºC for 15 min, and 95ºC for 2 min, followed by 45 cycles of 95ºC for 3 sec and 55ºC for 30 sec. Adequate internal controls, viral positive control and negative control were included during each run.

### Analytical sensitivity and reproducibility

The analytical sensitivity (i.e. minimum number of copies in a sample measured accurately), expressed by the limit of detection (LOD) of the assay was determined using six 10-fold serial dilutions of a synthetic plasmid of the full nucleocapsid gene according to datasheet information (2019-CoV Plasmid Control, IDT), from 1×10^5^ to 1 viral copies/µL in ultra-pure water. Dilutions were run in duplicate through both, dye-based and probe-based, assays. A standard curve was obtained plotting the C_T_ values against the copy number. The reaction efficiency was determined by the calculation of the correlation coefficient (*R*^2^). The repeatability and reproducibility of the assay was evaluated in four 10-fold serial dilutions of the standard plasmids (from 1×10^5^ to 1×10^2^) in duplicates using intra- and inter-assay tests within the same run and through two independent runs, respectively. The mean, standard deviations (SD), and coefficient of variation (CV) were calculated for each dilution based on their C_T_ values.

### Performance comparison of RT-qPCR assays

We compared the performance of the dye-based with the probe-based assay by analyzing 610 nasopharyngeal samples. Viral RNA was isolated as describe above. Each sample was individually analyzed using both methods in parallel, using adequate internal controls, viral positive control and negative control.

### Pooling test

In order to evaluate the ability of the RT-qPCR assay to scale for massive use, we run our tests in two different group systems: pooling after RNA extraction (post-RNA extraction) and pooling before RNA extraction (pre-RNA extraction) (Figure 3B and 3C). For the pooling post-RNA extraction, the eluted RNA from a confirmed positive sample of a patient was added to pools of 5, 10, 15 and 20 RNA negative samples. The positive sample was added at two different virus copy numbers emulating infected individuals at mid (∼400 copies/µL) and a high (∼6,000 copies/ µL) viral load (Kim et al 2020; Zou et al 2020). A total of 5 µL of RNA sample was added to the final RT-qPCR mix. Each pool was run in duplicate. An only negative pool were run within each assay. A positive plasmid control (PC) at 2,000 viral copies/µl was included. For the pooling pre-RNA extraction, we tested pools of 5 and 10 samples using both RT-qPCR assays. A 100 µL of viral transport media of each individual samples were mixed into a single tube. Then, a total of 200 µL of the mix was used for viral extraction, as described above, with a final elution of 60 µL. We compared adding volumes of 5 µL and 10 µL of RNA template added to the RT-qPCR mix. We evaluated 10 pools using a set of positive samples with C_T_ values from 14.76 to 38.2, emulating patients from a wide spectrum of viral load. We further compared 15 pools of 10 samples each adding 5 µL and 13.5 µL of RNA template, which is the maximum volumen allowed in a 20 µL RT-qPCR mix. All pools include one positive sample that with a C_T_ values that ranged from the 12.7 to 39.9. An only negative pool was run within each assay.

## RESULTS

### Evaluation of a dye-based assay

All set of primers targeting nucleocapsid gene amplified correctly using the dye-based real-time RT-PCR assay. One pair of primers (N1-F and N1-R) was selected for further efficiency and performance based on the highest sensitivity (lowest C_T_) obtained minimizing non-specific amplification products during melting curve analysis. The standard curve of the C_T_ against the amount of synthetic viral plasmid showed a wide dynamic range from 1 to 1×10^5^ viral copies/µL in 40 cycles, a correlation coefficient, *R*^2^, of 0.99 with a standard curve slope of –3.715, and a reaction efficiency, *E*, of 86% (Figure 1A). The limit of detection of SARS-CoV-2 using the dye-based assay was at a dilution of 10 viral copies/µL with a mean C_T_ value of 35.31. This value was 1.09 cycles higher than the probe-based assay at the same dilution (Figure 1B). Further, across all dilutions, the C_T_ difference among assays was almost zero (i.e. 0.08), ranging from –1.09 to 0.87 cycles (Table 2). The dissociation curve performed after completed RT-qPCR peaked at 83.51 ± 0.25 ºC (Figure 2). All internal controls amplified correctly. No positive fluorescence amplification signal was obtained from negative controls and nuclease-free water. Standard curves for each assay were highly reproducible with no significant differences in slopes among runs of the same assay. The intra-assay SD and CV ranged from 0.02 to 0.14 and 0.1 to 0.44%, respectively, whereas the inter-assay ranged from 0.04 to 0.18 and 0.05 to 0.43% (Table 3), indicating that the dye-based assay is highly repeatable and reproducible.

**Table 2.**
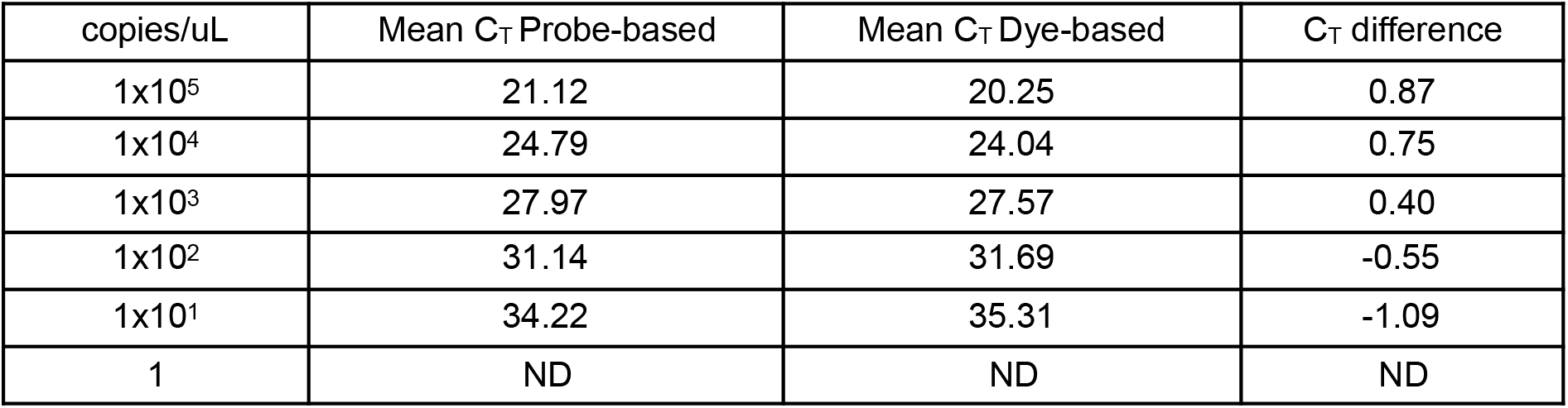
Cycles threshold (C_T_) obtained from six 10-fold dilutions using the probe-based and dye-based real-time RT-PCR assays. ND = Not detected.

| copies/uL | Mean $C_T$ Probe-based | Mean $C_T$ Dye-based | $C_T$ difference |
| --- | --- | --- | --- |
| 1x10<sup>5</sup> | 21.12 | 20.25 | 0.87 |
| 1x10<sup>4</sup> | 24.79 | 24.04 | 0.75 |
| 1x10<sup>3</sup> | 27.97 | 27.57 | 0.40 |
| 1x10<sup>2</sup> | 31.14 | 31.69 | -0.55 |
| 1x10<sup>1</sup> | 34.22 | 35.31 | -1.09 |
| 1 | ND | ND | ND |

**Table 3.** Intra-assay repeatability and inter-assay reproducibility for the dye-based real-time RTPCR assays.

| copies/uL | Intra-assay variability |  | Inter-assay variability |  |
| --- | --- | --- | --- | --- |
| | Mean ± SD | CV (%) | Mean ± SD | CV (%) |
| 1x10<sup>5</sup> | 20.25 ± 0.02 | 0.1 | 20.36 ± 0.16 | 0.05 |
| 1x10<sup>4</sup> | 24.04 ± 0.03 | 0.13 | 24.09 ± 0.07 | 0.15 |
| 1x10<sup>3</sup> | 27.57 ± 0.06 | 0.22 | 27.7 ± 0.18 | 0.17 |
| 1x10<sup>2</sup> | 31.69 ± 0.14 | 0.44 | 31.72 ± 0.04 | 0.43 |

**Figure 1.**
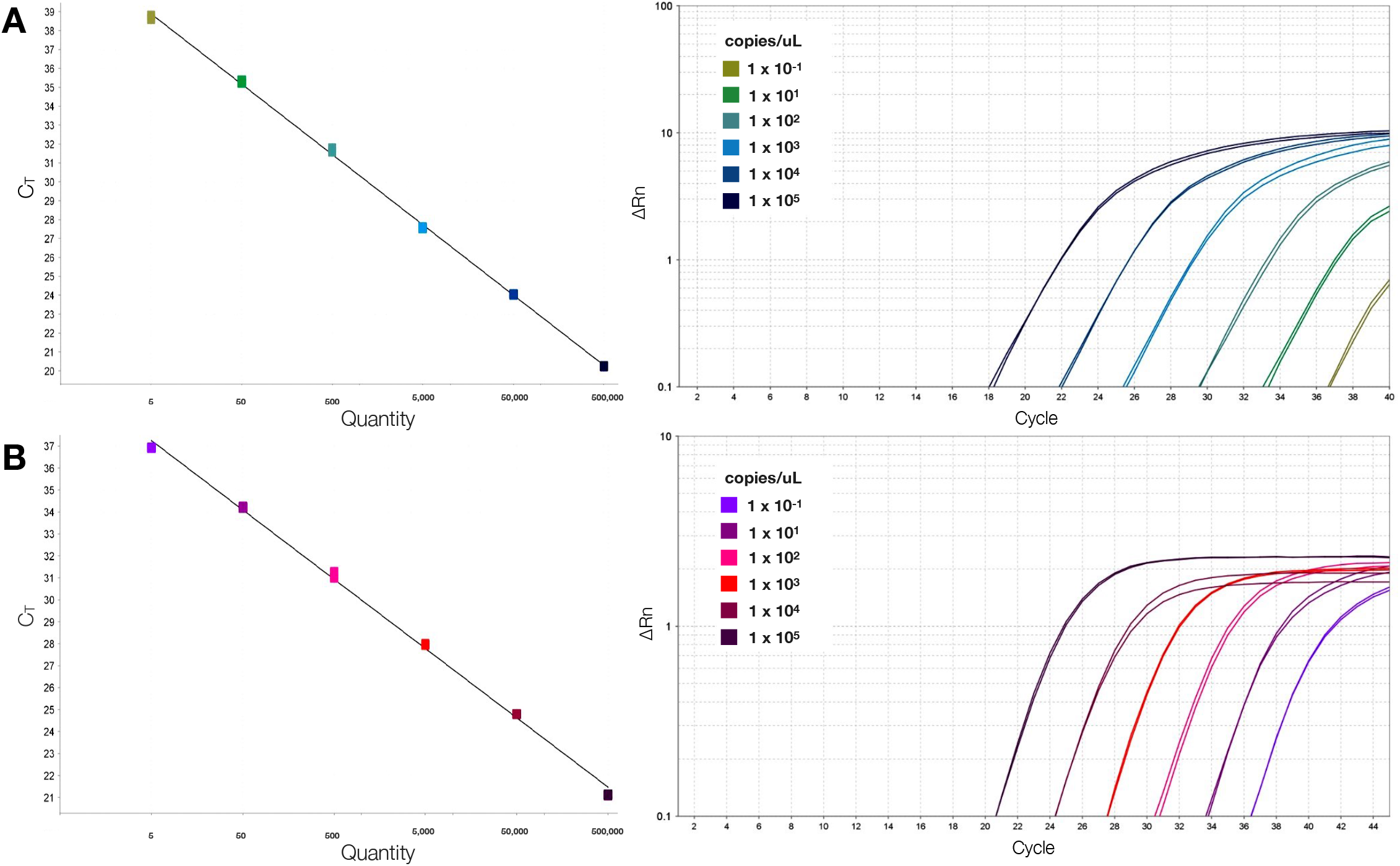
Amplification and standard curves of the SARS-CoV-2, obtained from 10-fold dilutions of the viral nucleocapsid plasmid by dye-based (A) and probe-based (B) RT-qPCR. Each point represents duplicate amplification at each dilution.

**Figure 2.**
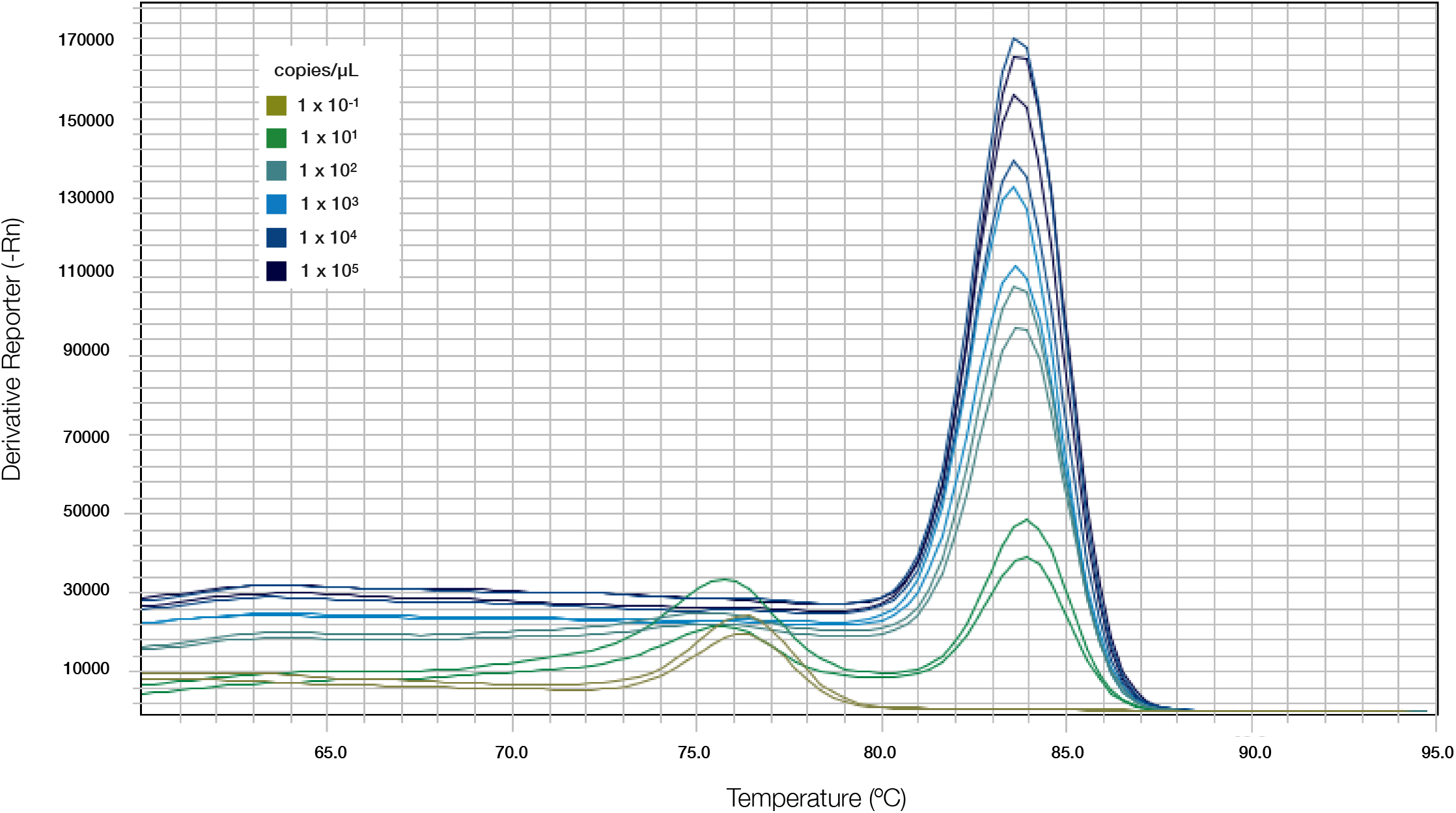
Fluorescence melting curve analysis for the detection of the nucleocapsid gene of SARS-CoV-2 of dye-based assay at six different n 10-fold dilutions.

### Performance comparison among assays

A total of 610 nasopharyngeal samples were examined by dye- and probe-based assays in order to compare performance among assays. The probe- and dye-based RT-qPCR detected 24 and 20 positive samples to SARS-CoV-2, respectively. The estimated sensitivity and specificity of the dye-based RT-qPCR was 83.3% (95% CI: 62.9–95.3%) and 100% (95% CI: 99.4–100%), respectively. In addition, positive samples from probe-based showed always a lower C_T_ value than dye-based assays (range: 0.72 to 5.9 cycles).

### Pooling Test

We compared the RT-qPCR performance of individual samples (Figure 3A) with pooled samples grouped at two different analytical stages: after (Figure 3B) and before (Figure 3C) the viral RNA extraction. In the post-RNA extraction pools, RNA of SARS-CoV-2 was detected in all pools, independent the amount of starting viral material, emulating patients with two different viral loads on each pool. All pools, in both dye- and probe-based, showed a negative impact on the performance of the RT-qPCR, increasing the C_T_ values when compared with individual testing (Figure 4). Using a high viral copies sample (∼6,000 copies/µL), the mean C_T_ values of pools ranged from 27.14 to 29.23 that were 4.12 to 6.22 cycles from the original sample using the dye-based assay, and from 23.84 to 25.68 at 2.59 to 4.43 cycles away from the original sample using the probe-based sample (Table 4). Similarly, pools emulating a patient with a mid-viral load (∼400 copies/µL) showed C_T_ values from 30.40 to 31.83 which were 3.10 to 4.53 cycles from the original sample in the dye-based assays, whereas using the probe-based assay the C_T_ values were less than 2 cycles from the original sample (Table 4). None of the negative pools by probe-based RT-qPCR showed fluorescence amplification signal. The same negative pools showed a high C_T_ value using the dye-based assay, although no melting products were detected at the 83°C target peak.

**Figure 3.**
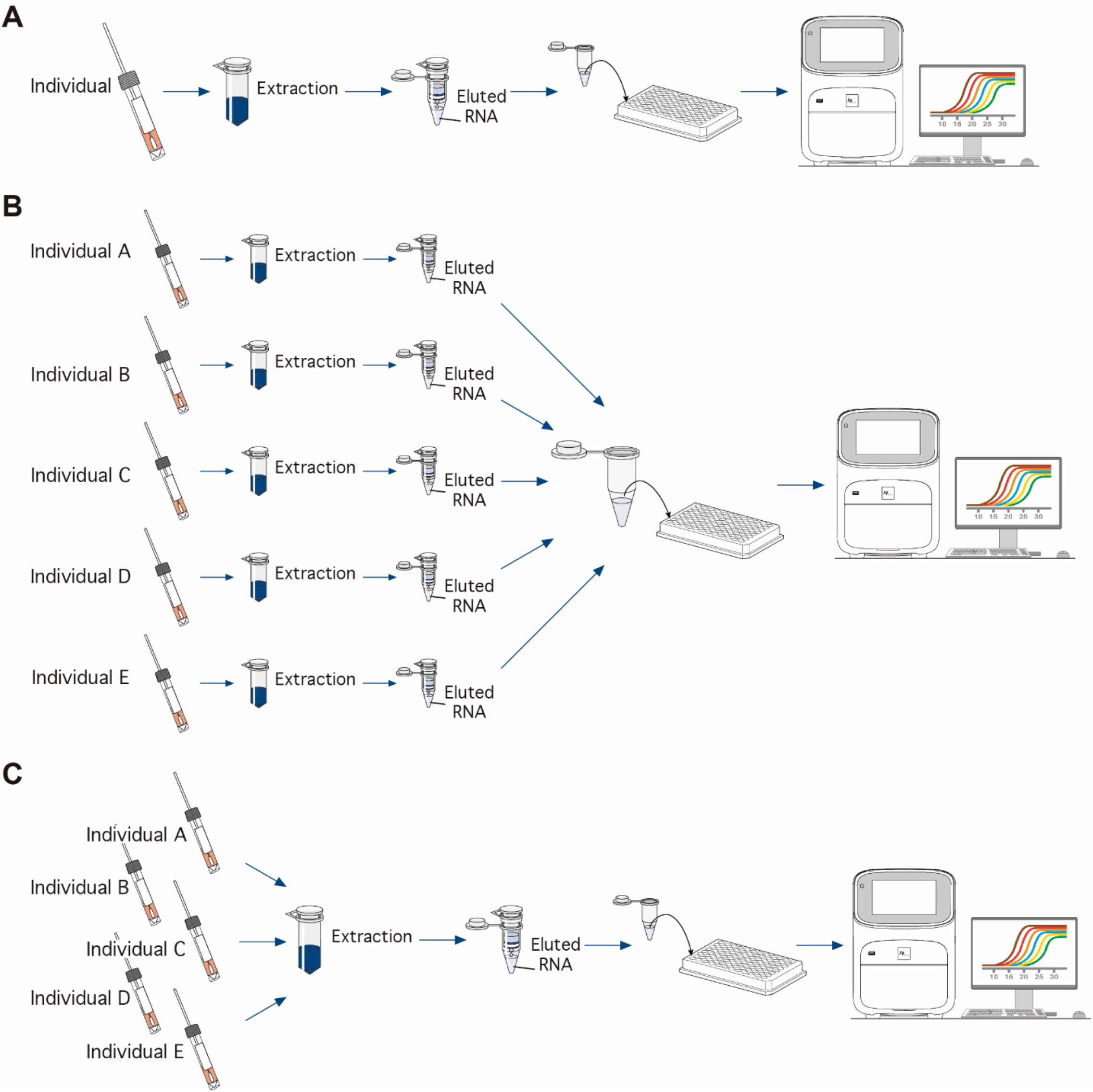
Schematic diagram of the experiments of individual (A), and pooling systems in a pre-RNA extraction (B) and post-RNA extraction (C).

**Figure 4.**
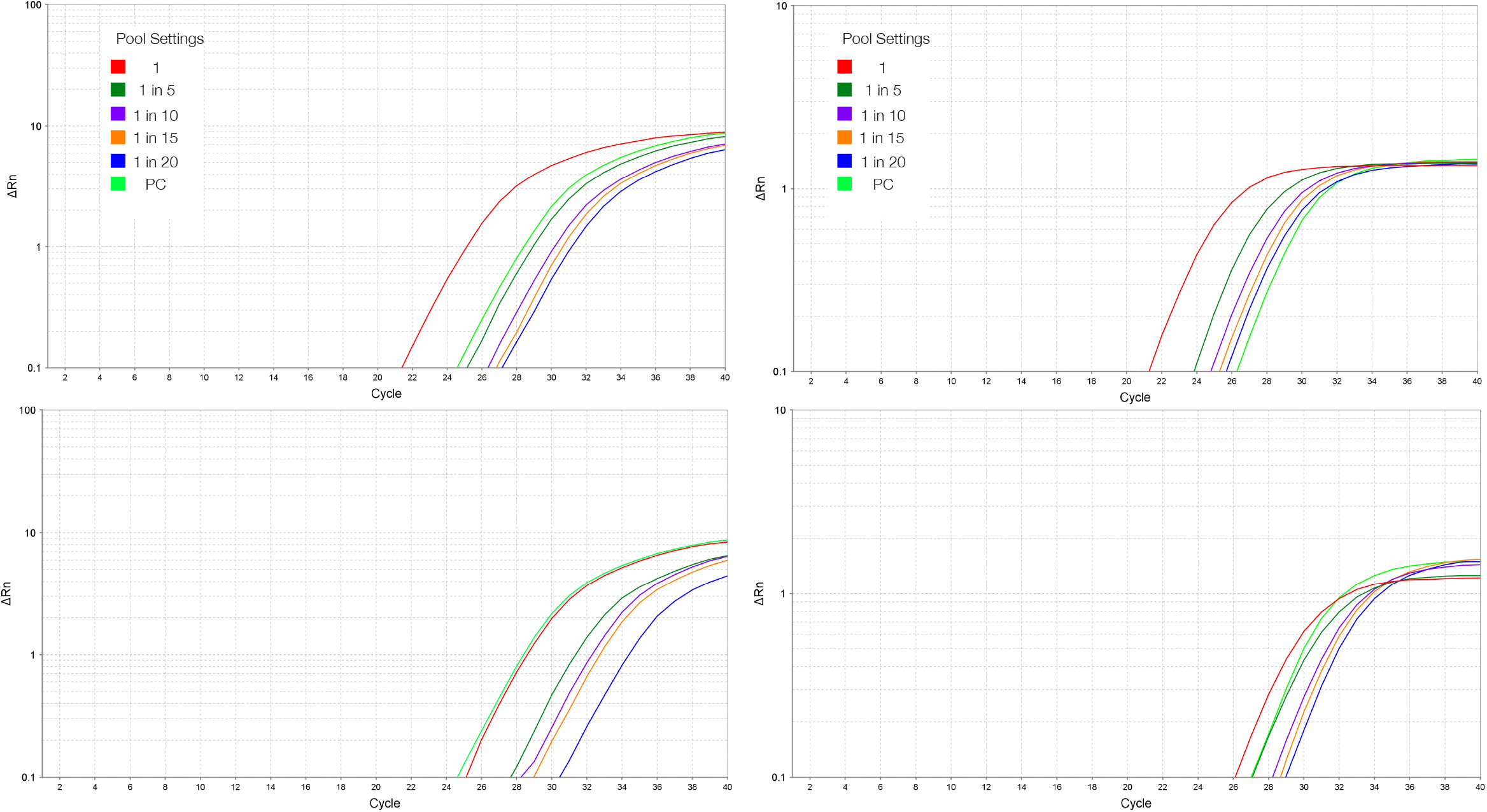
Amplification curves of SARS-CoV-2 sample in post-RNA extraction pool testing using 5, 10, 15 and 20 samples at mid- (above) and high (below) viral load sample using dye-based (left) and probe-based (right) assay.

**Table 4.** Cycles threshold (C_T_) obtained from four different pool post-RNA extraction settings using a probe-based (Probe) and dye-based (Dye) RT-qPCR assay. The original (positive) sample is denoted as 1.

| Pool Setting | High Viral Load |  |  |  | Low Viral Load |  |  |  |
| --- | --- | --- | --- | --- | --- | --- | --- | --- |
| | Probe Mean $C_T$ | $C_T$ Difference | Dye Mean $C_T$ | $C_T$ Difference | Probe Mean $C_T$ | $C_T$ Difference | Dye Mean $C_T$ | $C_T$ Difference |
| 1 | 21.25 | 0 | 23.32 | 0 | 26.12 | 0 | 27.30 | 0 |
| 1:5 | 23.84 | 2.59 | 27.14 | 3.82 | 27.09 | 0.97 | 30.40 | 3.10 |
| 1:10 | 24.79 | 3.55 | 28.42 | 5.1 | 28.12 | 0.89 | 31.17 | 3.87 |
| 1:15 | 25.32 | 4.1 | 28.98 | 5.66 | 28.65 | 1.36 | 31.69 | 4.39 |
| 1:20 | 25.68 | 4.43 | 29.23 | 5.91 | 28.98 | 1.69 | 31.83 | 4.53 |

In the pre-RNA extraction pools, we evaluated pools using individual positive samples with a wide range of individual C_T_ values. The probe-based assay was able to detect viral amplification in 8/10 pools of size 5 and 10, using either 5 or 10 µL of RNA template. In all pools, the C_T_ values increased when compared with individual testing. As expected, the C_T_ was greater when the pool size increased from 5 (C_T_ mean difference of 2.83) to 10 (C_T_ mean difference of 7.1) (Table 5). When the amount of RT-qPCR template was doubled from 5 to 10 µL, the C_T_ difference with the individual sample decreased (Figure 5A and B). We were able to detect pools that contained an individual positive sample with C_T_ value up to 35. The two pools containing samples with the highest individual C_T_ values, 37.96 and 38.2, were not detected by pooling in either 5 or 10 and adding 5 or 10 µL. When the volume of RNA template increased from 5 to 13.5 µL, all pools of 10 samples (15/15) containing individual positive samples with C_T_ values from 12.7 to 39.9 were detected. In contrast, 13/15 pools adding 5 µL of RNA template were detected, where two pools with individual positive sample C_T_ of 33.5 and 39.9 were not detected (Table 6). All C_T_ values decreased when the volume of RNA template increased (Figure 5C). The performance of the dye-based assay in pre-RNA extraction pools resulted in the amplification of 6/11 and 5/11 in pools of size 5 and 10, respectively, using 5 µL of RNA template (Table 6). Surprisingly, increasing the amount of RNA template to 10 µL did not increase the detection rate. In fact, adding more template to the RT-qPCR mix, the detection decrease in 3/11 and 0/11 in pools of 5 and 10, respectively,

**Table 5.** Effect of increasing the pool size in the RT-qPCR performance. Cycles threshold (C_T_) obtained from pools of 5 and 10 pre-RNA extraction samples using 10 uL of RNA template in a probe-based RT-qPCR assay. The original sample is denoted as individual C_T_. _CT_ difference corresponds to pool C_T_ value minus individual C_T_. ND = Not detected.

| ID | Individual $C_T$ | Pool 1:5 $C_T$ value | $C_T$ Difference | Pool 1:10 $C_T$ value | $C_T$ Difference |
| --- | --- | --- | --- | --- | --- |
| Pool1 | 14.76 | 19.13 | 4.37 | 25.47 | 10.71 |
| Pool2 | 18.40 | 22.99 | 4.59 | 28.66 | 10.26 |
| Pool3 | 19.82 | 22.99 | 3.17 | 27.73 | 7.92 |
| Pool4 | 22.01 | 25.47 | 3.47 | 33.54 | 11.53 |
| Pool5 | 23.01 | 27.08 | 4.07 | 32.17 | 9.16 |
| Pool6 | 26.62 | 27.77 | 1.15 | 29.83 | 3.21 |
| Pool7 | 27.62 | 28.68 | 1.06 | 29.3 | 1.68 |
| Pool8 | 31.09 | 31.82 | 0.73 | 33.43 | 2.34 |
| Pool9 | 37.96 | ND | ND | ND | ND |
| Pool10 | 38.20 | ND | ND | ND | ND |
| | | Mean $C_T$ value: | 2.83 | Mean $C_T$ value: | 7.10 |

**Table 6.** Effect of increasing the volume of RNA template in the probe-based RT-qPCR performance. Cycles threshold (C_T_) obtained from pools of 10 pre-RNA extraction samples using 5 and 13.5 uL of RNA template in a probe-based RT-qPCR assay. The original sample is denoted as individual C_T_. C_T_ difference corresponds to pool C_T_ value minus individual C_T_. ND = Not detected.

| ID | Individual $C_T$ | 5 μL Pool $C_T$ value | $C_T$ Difference | 13.5 μL Pool $C_T$ value | $C_T$ Difference |
| --- | --- | --- | --- | --- | --- |
| Pool 11 | 12.69 | 15.65 | 2.96 | 14.2 | 1.51 |
| Pool 12 | 12.86 | 15.13 | 2.27 | 13.8 | 0.94 |
| Pool 13 | 14.76 | 26.34 | 11.59 | 23.48 | 8.72 |
| Pool 14 | 18.40 | 29.82 | 11.42 | 27.30 | 8.90 |
| Pool 15 | 19.82 | 29.26 | 9.44 | 26.60 | 6.78 |
| Pool 16 | 22.01 | 35.46 | 13.45 | 31.99 | 9.98 |
| Pool 17 | 23.01 | 33.69 | 10.68 | 31.24 | 8.23 |
| Pool 18 | 26.6 | 28.02 | 1.42 | 26.76 | 0.16 |
| Pool 19 | 26.62 | 31.82 | 5.20 | 28.65 | 2.03 |
| Pool 20 | 30.04 | 31.52 | 1.48 | 30.25 | 0.21 |
| Pool 21 | 31.09 | 35.40 | 4.31 | 31.86 | 0.77 |
| Pool 22 | 31.87 | 33.96 | 2.09 | 32.44 | 0.57 |
| Pool 23 | 33.48 | ND | ND | 38.59 | 5.11 |
| Pool 24 | 34.67 | 39.62 | 4.94 | 38.43 | 3.76 |
| Pool 25 | 39.9 | ND | ND | 38 | -1.90 |
| | | Mean $C_T$ value: | 6.25 | Mean $C_T$ value: | 3.72 |

**Table 7.** Cycles threshold (C_T_) obtained from pools of 5 and 10 pre-RNA extraction samples using 5 and 10 uL of RNA template in a dye-based RT-qPCR assay. The original sample is denoted as individual C_T_. ND = Pools with not detectable amplification signal or melting at non-expected temperature.

| ID | Individual $C_T$ | 5 μL RNA template | | 10 μL RNA template | |
| --- | --- | --- | --- | --- | --- |
| | | Pool 1:5 $C_T$ value | Pool 1:10 $C_T$ value | Pool 1:5 $C_T$ value | Pool 1:10 $C_T$ value |
| Pool 1 | 17.95 | 27.43 | 36.46 | 34.38 | ND |
| Pool 2 | 22.24 | 29.66 | 35.77 | 32.42 | ND |
| Pool 3 | 23.33 | 30.10 | 39.02 | ND | ND |
| Pool 4 | 26.27 | 31.54 | 38.52 | ND | ND |
| Pool 5 | 26.98 | 34.88 | ND | ND | ND |
| Pool 6 | 31.84 | 34.60 | 38.59 | 38.06 | ND |
| Pool 7 | 34.42 | ND | ND | ND | ND |
| Pool 8 | 36.08 | ND | ND | ND | ND |
| Pool 9 | 36.89 | ND | ND | ND | ND |
| Pool 10 | 38.66 | ND | ND | ND | ND |
| Pool 11 | 39.20 | ND | ND | ND | ND |

**Figure 5.**
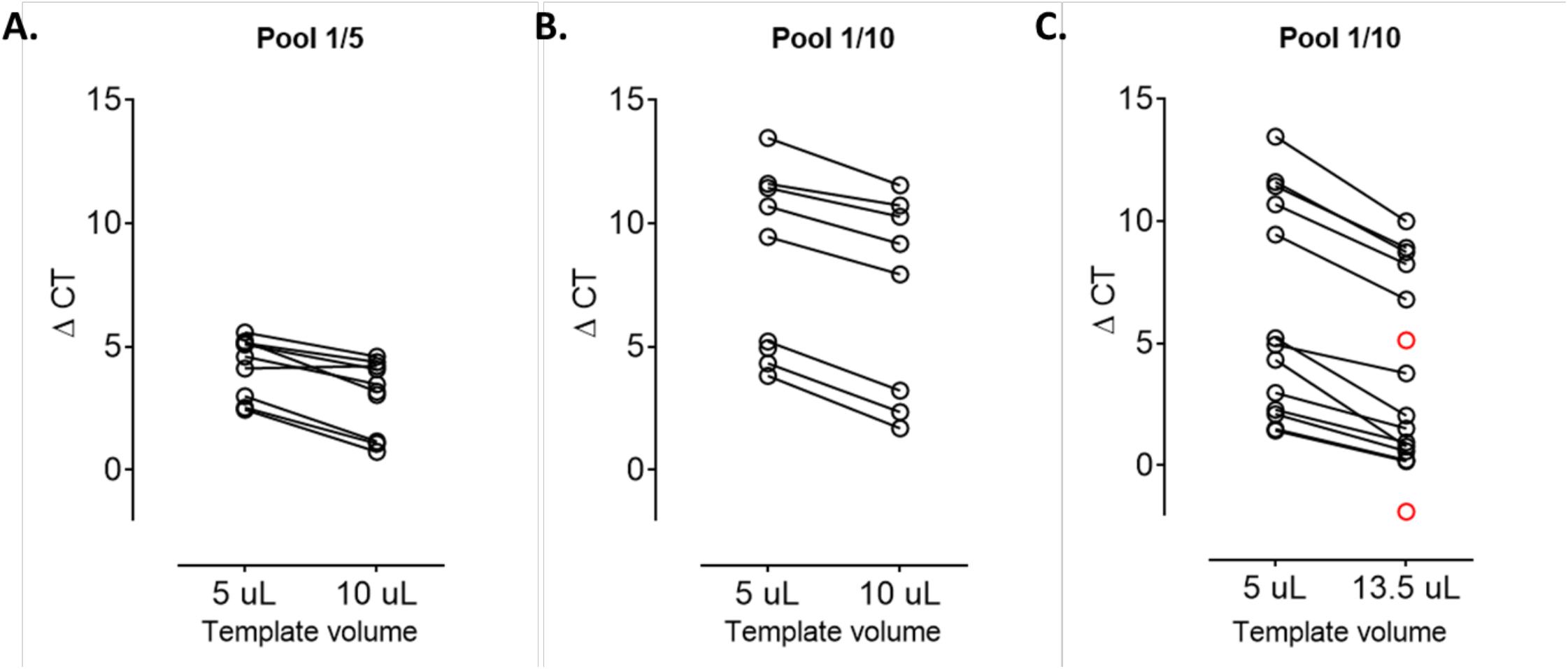
Cycles threshold (C_T_) difference (∆C_T_) using 5 µl and 10 µl of RNA template in pre-RNA extraction pool testing of 5 (A) and 10 (B) samples (n = 10). (C) ∆CT using 5 µl and 13.5 µl of RNA template in pre-RNA extraction pool testing of 10 samples (n = 15). Samples with no coupled dot are denoted in red.

## DISCUSSION

In this study, we compared a dye-based and probe-based real-time RT-qPCR assay for the economic and rapid detection and quantification of SARS-CoV-2 in human samples by individual and pooling testing. The dye-based technique showed a high analytical sensitivity similar to the probe-based detection assay used worldwide. Further, we showed that this assay may also be applicable in testing by pool systems post-RNA extraction, whereas the probe-based assay have a excellent performance in pre-RNA extraction pooling tests. Thus, each test has particular advantages that can be exploited for massive use to increase sensitivity in surveillance programs in large target populations.

Our results showed that using the dye-based assay describe here, the SARS-CoV-2 can be detected up to 50 viral copies (a dilution of 10 copies/µL) of the RNA template, showing a similar analytical sensitivity obtained with the probe-based standard technique (Table 2). The LODs described here are within the range reported for COVID-19 detection in two comparative studies using up to six approved RT-qPCR kit assays for SARS-CoV-2 (van Kasteren et al, 2020; Wang et al, 2020) and similar for other human coronaviruses (Goka et al. 2014, Adachi et al 2004). We further evaluated this method in 610 nasopharyngeal samples, showing that the individual test of the dye-based assay had a similar performance to the probe-based assay in detecting positive samples. An amplification curve might be detectable at high C_T_ values in negative individuals. Thus, a critical step in the proper use and interpretation of the dye-based technique is the analysis of the melting curve reducing the chances of obtaining false positives. We encourage the use of this test for individual surveillance testing when group sensitivity is a priority.

The pooling or group testing can be a very efficient and effective methodology for the implementation of massive screening in surveillance programs, reducing costs, laboratory time, and the total number of reactions applied. Recently, several pooling tests for COVID-19 using probe-based assays have shown efficient viral detection. These studies have shown a wide range of pooling settings, from eight to 32 samples per pool, increasing up to 8 folds the testing efficiency compared to individual testing (Shenta et al, 2020; Viehweger et al, 2020; Ben-Ami et al, 2020). Here, we showed an efficient performance in the range of setting pools tested before the RNA extraction step, with an analytical sensitivity that shown that pooling setting between 5 to 20 samples per pool retain the accuracy of the test obtaining a shift to the right of the amplification curve up to 5.9 cycles in the dye-based assay and 4.4 cycles in the probe-based assay. The C_T_ values obtained from group testing are highly dependent on the viral load of the positive sample in the pool and the number of samples included in the pool setting. Thus, a compromise of C_T_ values is expected when samples are pooled and even more in samples with mid or low viral load. This compromise in cycles was present in both probe- and dye-based assays. However, an increase of C_T_ values from 33,1 to 35,6 cycles (2.5 cycles) has been shown between individual analysis and pool test with 8 samples (1/7 positive), respectively (Gupta et al, 2020). Similarly, another study showed that a positive sample of SARS-CoV-2 can be detected using TaqMan assays in pooled samples without compromising the assay sensitivity (Ben-Ami et al, 2020). Further, a study using samples with high or low viral loads, showed no significant differences in C_T_ values between pooled and individual samples, suggesting that sensitivity was not affected by pooling specimens, regardless of the viral load (Wacharapluesadee et al, 2020). We further explored the use of pooling samples previous to the analytical stage of RNA extraction. This is intended to significantly save time, laboratory staff effort, and economic resources in a surveillance program. Using the probe-base assay, we were able to detect positive individuals with C_T_ values of 31.09 in pools of 5 and 10 samples using 5 or 10 µL of RNA template. Pooled samples with one individual with C_T_ over 38 were not detected. Due to the lack of samples between C_T_ 32 and 38, we cannot rule out the analytical sensitivity of this assay at C_T_ 32 using 10 µL of RNA template but further analyses might be pursued to detect a more trustful detection threshold. Since increasing the amount of the RNA template from 5 to 10 µL we observed less compromise of cycles, we further explore increasing to the maximum amount of RNA template allowed in a 20 µL RT-qPCR mix, with a set of positive samples with a wider range of C_T_ values (12.7 to 39.9). When the template volume increased to 13.5 µL in pools of 10, we were able to reduce the delta C_T_ from 6.25 (using 5 µL of RNA template) to 3.72 on average and increase the detection rate to 100%.

Contrary to our expectations, we observed an increase in the C_T_ value the RNA template volume increased using the dye-based assay. The increase in C_T_ value when using 10 µL versus 5 µL of RNA template in the dye-based RT-qPCR assay could be explained by the effect of inhibitors present in the extracted RNA sample, although this effect was not observed in the reaction with the probe-based assay. This effect might be related to differences in the sensitivity to inhibitors of the Transcriptase reverse and/or Taq Polymerase included on each master mixes (Levesque-Sergerie et al., 2007). The presence of inhibitors of the RT-qPCR reaction is a recurrent problem in clinical samples and the effect on the result of the reaction will depend on the enzymes (i.e. Transcriptase Reverse (M-MLV) and DNA Polymerase (Taq Polymerase)) used (Schrader et al., 2012). Since the dye-based assay was not able to perform adequately using this approach, we do not recommend the use of this protocol in pooled pre-RNA extraction samples.

For massive surveillance use, we are confident to suggest the use of the probe-based assay in a pre-RNA extraction pooling testing up to 10 samples. For a system of pooling post-RNA extraction, we suggest first the use of probe-based assay followed by dye-based assays, and both can be used in pools up to 20 samples. Implementing a pool system for population sampling results in an important savings of laboratory resources and time, which are two key factors during an epidemic outbreak, such as for COVID-19, that may limit the surveillance approach selected for viral detection testing in the population. Further, the individual testing use, the dye-based performed similarly to the probe-based assays and could be used in case of a shortage of probe resources.

COVID-19 is a highly contagious disease causing devastating effects worldwide. Effective interventions are mandatory to control the transmission and spread of the virus. Using the pooling approaches evaluated here, we are confident that it can be used as a valid alternative assay for the detection of SARS-CoV-2 in human samples. The low cost and the potential use in pre-RNA extraction pool systems, place of this assays as a valuable resource for scalable sampling to larger populations such as surveillance targeting asymptomatic and presymptomatic individuals where massive testing is essential for the rapid identification of potential spreaders.

## Data Availability

The authors confirm that the data supporting the findings of this study are available within the article, and/or on request from the corresponding author, CV.

## ACKNOWLEDGEMENTS

We thanks Gobierno Regional de Los Rios and Universidad Austral de Chile for the financial support. Also, we thanks Dr. Marcela Perez (Hospital Lanco), Mr. Luis Leyton and Dr. Maritza Navarrete (Hospital Base de Valdivia), Dr. Omar Ulloa (Ejército de Chile), and Dr. Claudio Henríquez (Universidad Austral de Chile) for logistic and technical support.

## REFERENCES

Adachi, D., Johnson, G., Draker, R., Ayers, M., Mazzulli, T., Talbot, P.J., Tellier, R., 2004. Comprehensive detection and identification of human coronaviruses, including the SARS-associated coronavirus, with a single RT-PCR assay. Journal of Virology Methods 122, 29–36.

Andersen, K.G., Rambaut, A., Lipkin, W.I., Holmes, E.C., Garry, R.F., 2020. The proximal origin of SARS-CoV-2. Nature Medicine 26, 450–452.

Bai, Y., Yao, L., Wei, T., Tian, F., Jin, D.-Y., Chen, L., Wang, M., 2020. Presumed Asymptomatic Carrier Transmission of COVID-19. Journal of the American Medical Association 323, 1406–1407.

Ben-Ami, R., Klochendler, A., Seidel, M., Sido, T., Gurel-Gurevich, O., Yassour, M., Meshorer, E., Benedek, G., Fogel, I., Oiknine-Djian, E., Gertler, A., Rotstein, Z., Lavi, B., Dor, Y., Wolf, D.G., Salton, M., Drier, Y., 2020. Pooled RNA extraction and PCR assay for efficient SARS-CoV-2 detection. medRxiv. https://doi.org/10.1101/2020.04.17.20069062

CDC, 2020. Coronavirus Disease 2019 (COVID-19).

Eis-Huebinger, A.M., Hoenemann, M., Wenzel, J.J., Berger, A., Widera, M., Schmidt, B., Aldabbagh, S., Marx, B., Streeck, H., Ciesek, S., Liebert, U.G., Huzly, D., Hengel, H., Panning, M., 2020. Ad hoc laboratory-based surveillance of SARS-CoV-2 by real-time RTPCR using minipools of RNA prepared from routine respiratory samples. medRxiv. https://doi.org/10.1101/2020.03.30.20043513

Foddai, A., Lindberg, A., Lubroth, J., Ellis-Iversen, J., 2020. Surveillance to improve evidence for community control decisions during the COVID-19 pandemic – Opening the animal epidemic toolbox for public health. One Health. (Amsterdam, Netherlands). https://doi.org/10.1016/j.onehlt.2020.100130

Goka, E.A., Vallely, P.J., Mutton, K.J., Klapper, P.E., 2015. Pan-human coronavirus and human bocavirus SYBR Green and TaqMan PCR assays; use in studying influenza A viruses coinfection and risk of hospitalization. Infection 43(2), 185–192.

Gupta, E., Padhi, A., Khodare, A., Aggarwal, R., Ramachandran, K., Mehta, V., Kilikdar, M., Dubey, S., Kumar, G., Sarin, S.K., 2020. Pooled RNA sample reverse transcriptase real time PCR assay for SARS CoV-2 infection: a reliable, faster and economical method. medRxiv. https://doi.org/10.1101/2020.04.25.20079095

He, X., Lau, E.H.Y., Wu, P., Deng, X., Wang, J., Hao, X., Lau, Y.C., Wong, J.Y., Guan, Y., Tan, X., Mo, X., Chen, Y., Liao, B., Chen, W., Hu, F., Zhang, Q., Zhong, M., Wu, Y., Zhao, L., Zhang, F., Cowling, B.J., Li, F., Leung, G.M., 2020. Temporal dynamics in viral shedding and transmissibility of COVID-19. Nature Medicine. https://doi.org/10.1038/s41591-020-0869-5

Hellewell, J., Abbott, S., Gimma, A., Bosse, N.I., Jarvis, C.I., Russell, T.W., Munday, J.D., Kucharski, A.J., Edmunds, W.J., Sun, F., Flasche, S., Quilty, B.J., Davies, N., Liu, Y., Clifford, S., Klepac, P., Jit, M., Diamond, C., Gibbs, H., Zandvoort], K. [van, Funk, S., Eggo, R.M., 2020. Feasibility of controlling COVID-19 outbreaks by isolation of cases and contacts. Lancet Global Health 8, e488–e496. https://doi.org/10.1016/S2214-109X(20)30074-7

Kim JY, Ko JH, Kim Y, Kim YJ, Kim JM, Chung YS, Kim HM, Han MG, Kim SY, Chin BS. Viral Load Kinetics of SARS-CoV-2 Infection in First Two Patients in Korea. J Korean Med Sci. 2020 Feb 24;35(7):e86. https://doi.org/10.3346/jkms.2020.35.e86

Levesque-Sergerie, J., Duquette, M., Thibault, C. Delbecchi, L., Bissonnette N., 2007 Detection limits of several commercial reverse transcriptase enzymes: impact on the low-and high-abundance transcript levels assessed by quantitative RT-PCR. BMC Molecular Biology 2007, 8:93.

Li, R., Pei, S., Chen, B., Song, Y., Zhang, T., Yang, W., Shaman, J., 2020. Substantial undocumented infection facilitates the rapid dissemination of novel coronavirus (SARSCoV2). Science 368, 489–493.

Li, W., Zhang, C., Sui, J., Kuhn, J.H., Moore, M.J., Luo, S., Wong, S.-K., Huang, I.-C., Xu, K., Vasilieva, N., Murakami, A., He, Y., Marasco, W.A., Guan, Y., Choe, H., Farzan, M., 2005. Receptor and viral determinants of SARS-coronavirus adaptation to human ACE2. The EMBO Journal 24, 1634–1643.

Liu, Y., Gayle, A.A., Wilder-Smith, A., Rocklöv, J., 2020. The reproductive number of COVID-19 is higher compared to SARS coronavirus. Journal of Travel Medicine 27. https://doi.org/10.1093/jtm/taaa021

Mizumoto, K., Kagaya, K., Zarebski, A., Chowell, G., 2020. Estimating the asymptomatic proportion of coronavirus disease 2019 (COVID-19) cases on board the Diamond Princess cruise ship, Yokohama, Japan, 2020. Eurosurveillance 25. https://doi.org/10.2807/1560-7917.ES.2020.25.10.2000180

Narayanan, K., Frost, I., Heidarzadeh, A., Tseng, K.K., Banerjee, S., John, J., Laxminarayan, R., 2020. Pooling RT-PCR or NGS samples has the potential to cost-effectively generate estimates of COVID-19 prevalence in resource limited environments. medRxiv. https://doi.org/10.1101/2020.04.03.20051995

Pareek, M., Greenaway, C., Noori, T., Munoz, J., Zenner, D., 2016. The impact of migration on tuberculosis epidemiology and control in high-income countries: a review. BMC Medicine 14, 48.

Peto, J., 2020. Covid-19 mass testing facilities could end the epidemic rapidly. BMJ 368, m1163. https://doi.org/10.1136/bmj.m1163

Peto, J., Alwan, N.A., Godfrey, K.M., Burgess, R.A., Hunter, D.J., Riboli, E., Romer, P., Buchan, I., Colbourn, T., Costelloe, C., Davey Smith, G., Elliott, P., Ezzati, M., Gilbert, R., Gilthorpe, M.S., Foy, R., Houlston, R., Inskip, H., Lawlor, D.A., Martineau, A.R., McGrath, N., McCoy, D., Mckee, M., McPherson, K., Orcutt, M., Pankhania, B., Pearce, N., Peto, R., Phillips, A., Rahi, J., Roderick, P., Saxena, S., Wilson, A., Yao, G.L., 2020. Universal weekly testing as the UK COVID-19 lockdown exit strategy. The Lancet 395 (10234), 1420–1421.

Prins, H.A.B., Verbon, A., Boucher, C.A.B., Rokx, C., 2017. Ending the epidemic: Critical role of primary HIV infection. The Netherlands Journal of Medicine 75, 321–327.

Ren, L.L., Wang, Y.M., Wu, Z.Q., Xiang, Z.C., Guo, L., Xu, T., Jiang, Y.Z., Xiong, Y., Li, Y.J., Li, X.W., Li, H., Fan, G.H., Gu, X.Y., Xiao, Y., Gao, H., Xu, J.Y., Yang, F., Wang, X.M., Wu, C., Chen, L., Liu, Y.W., Liu, B., Yang, J., Wang, X.R., Dong, J., Li, L., Huang, C.L., Zhao, J.P., Hu, Y., Cheng, Z.S., Liu, L.L., Qian, Z.H., Qin, C., Jin, Q., Cao, B., Wang, J.W., 2020. Identification of a novel coronavirus causing severe pneumonia in human: a descriptive study. Chinese Medical Journal 133(9), 1015–1024.

Shental, N., Levy, S., Skorniakov, S., Wuvshet, V., Shemer-Avni, Y., Porgador, A., Hertz, T., 2020. Efficient high throughput SARS-CoV-2 testing to detect asymptomatic carriers. medRxiv. https://www.medrxiv.org/content/10.1101/2020.04.14.20064618v1.

Schrader, C., Schielke, A., Ellerbroek, L. and Johne, R., 2012. PCR inhibitors – occurrence, properties and removal. J Appl Microbiol, 113: 1014–1026.

Tajadini, M., Panjehpour, M., Javanmard, S.H., 2014. Comparison of SYBR Green and TaqMan methods in quantitative real-time polymerase chain reaction analysis of four adenosine receptor subtypes. Advanced Biomedical Research 3, 85.

van Kasteren, P.B., van der Veer, B., van den Brink, S., Wijsman, L., de Jonge, J., van den Brandt, A.M., Molenkamp, R., Reusken, C.B.E.M., Meijer, A., 2020. Comparison of commercial RT-PCR diagnostic kits for COVID-19. bioRxiv. https://doi.org/10.1101/2020.04.22.056747

Viehweger, A., Kühnl, F., Brandt, C., König, B., 2020. Increased PCR screening capacity using a multi-replicate pooling scheme. medRxiv. https://doi.org/10.1101/2020.04.16.20067603

Wacharapluesadee, S., Kaewpom, T., Ampoot, W., Ghai, S., Khamhang, W., Worachotsueptrakun, K., Wanthong, P., Nopvichai, C., Supharatpariyakorn, T., Putcharoen, O., Paitoonpong, L., Suwanpimolkul, G., Jantarabenjakul, W., Hemachudha, P., Krichphiphat, A., Buathong, R., Plipat, T., Hemachudha, T., 2020. Evaluating efficiency of pooling specimens for PCR-based detection of COVID-19. medRxiv. https://doi.org/10.1101/2020.05.02.20087221

Walls, A.C., Park, Y.-J., Tortorici, M.A., Wall, A., McGuire, A.T., Veesler, D., 2020. Structure, Function, and Antigenicity of the SARS-CoV-2 Spike Glycoprotein. Cell 181, 281–292.e6.

Wang, X., Yao, H., Xu, X., Zhang, P., Zhang, M., Shao, J., Xiao, Y., Wang, H., 2020. Limits of Detection of Six Approved RT-PCR Kits for the Novel SARS-coronavirus-2 (SARS-CoV-2). Clinical Chemistry. https://doi.org/10.1093/clinchem/hvaa099

WHO, 2020a. Coronavirus disease 2019 (COVID-19) Situation Report – 90. World health Organization (WHO).

WHO, 2020b. Q&A: Similarities and differences – COVID-19 and influenza. World Health Organization.

Won, J., Lee, S., Park, M., Kim, T. Y., Park, M. G., Choi, B. Y., Kim, D., Chang, H., Kim, V. N., & Lee, C. J. (2020). Development of a Laboratory-safe and Low-cost Detection Protocol for SARS-CoV-2 of the Coronavirus Disease 2019 (COVID-19). Experimental neurobiology, 29(2), 107–119.

Wu, Z., McGoogan, J.M., 2020. Characteristics of and Important Lessons from the Coronavirus Disease 2019 (COVID-19) Outbreak in China: Summary of a Report of 72314 Cases From the Chinese Center for Disease Control and Prevention. Journal of the American Medical Association 323(13), 1239–1242.

Zou L, Ruan F, Huang M, Liang L, Huang H, Hong Z, et al. SARS-CoV-2 Viral Load in Upper Respiratory Specimens of Infected Patients. N Engl J Med. 2020;41(2):NEJMc2001737. https://doi.org/10.1056/NEJMc2001737

